# Addressing anti-syncytin antibody levels, and fertility and breastfeeding concerns, following BNT162B2 COVID-19 mRNA vaccination

**DOI:** 10.1101/2021.05.23.21257686

**Authors:** Citra NZ Mattar, Winston Koh, Yiqi Seow, Shawn Hoon, Aparna Venkatesh, Pradip Dashraath, Li Min Lim, Judith Ong, Rachel Lee, Nuryanti Johana, Julie SL Yeo, David Chong, Lay-Kok Tan, Jerry Chan, Mahesh Choolani, Paul Anantharajah Tambyah

## Abstract

**Objective:** To determine whether antibodies against the SARS-CoV-2 spike protein following BNT162B2 (Pfizer-BioNTech) COVID-19 mRNA vaccination cross-react with human syncytin-1 protein, and if BNT162B2 mRNA enters breast milk.

**Methods:** In this observational cohort study of female front-line workers with no history of COVID-19 infection, we amplified BNT162B2 mRNA in plasma and breast milk and assayed anti-SARS-CoV-2 neutralising antibodies and anti-human syncytin-1 binding antibodies in plasma, at early (1-4 days) and late (4-7 weeks) time points following first-dose vaccination.

**Results:** Fifteen consented participants (mean age 40.4 years, various ethnicities) who received at least one dose of BNT162B2, including five breast-feeding women and two women who were inadvertently vaccinated in early pregnancy, were recruited. BNT162B2 mRNA, detected by amplifying part of the spike-encoding region, was detected in plasma 1-4 days following the first dose (n=13), but not 4-5 weeks later (n=2), nor was the mRNA isolated from aqueous or lipid breast milk fractions collected 0-7 days post-vaccination (n=5). Vaccine recipients demonstrated strong SARS-CoV-2 neutralising activity by at least four weeks after the first dose (n=15), including the two pregnant women. None had placental anti-syncytin-1 binding antibodies at either time-point following vaccination.

**Conclusions:** BNT162B2-vaccinated women did not transmit vaccine mRNA to breast milk, and did not produce a concurrent humoral response to syncytin-1, suggesting that cross-reactivity to syncytin-1 on the developing trophoblast, or other adverse effects in the breast-fed infant from vaccine mRNA ingestion, are unlikely.

**What are the novel findings of this work?:** COVID-19 vaccination with BNT162B2 did not elicit a cross-reacting humoral response to human syncytin-1 despite robust neutralising activity to the SARS-CoV2 spike protein, and while vaccine mRNA was isolated from plasma, it was not found in breast milk.

**What are the clinical implications of this work?:** Our work directly addresses the fertility and breastfeeding concerns fuelling vaccine hesitancy among reproductive-age women, by suggesting that BNT162B2 vaccination is unlikely to cause adverse effects on the developing trophoblast, via cross-reacting anti-syncytin-1 antibodies, or to the breastfed neonate, via mRNA breast milk transmission.

## Introduction

Vaccine hesitancy threatens to compromise global vaccination efforts with lipid nanoparticle-encapsulated mRNA-based COVID-19 vaccines employed under emergency use authorisation to end the pandemic caused by the severe acute respiratory syndrome coronavirus 2 (SARS-CoV-2) and its emerging variants. Conspiracy theories propagated through social media cast doubts over the safety of these novel vaccines for fertility and breastfeeding,^1, 2^ such that 13% of unvaccinated persons in the USA believe that COVID-19 vaccination may result in infertility.^3^ One such claim generating significant traction suggests that vaccine mRNA-translated antibodies – which target the SARS-CoV-2 spike protein – cross-react with human syncytin-1, resulting in infertility and pregnancy loss. Syncytin proteins, which originate from endogenous human retroviruses, are involved in gamete fusion during fertilization and normal placental development,^4-8^ and amino acid sequence similarities between syncytin proteins and the SARS-CoV-2 spike protein S2 domain raise the possibility of cross-reacting antibodies following COVID-19 vaccination.^9^

This worrying spectre of disinformation may extend to BNT162B2 mRNA persistence, with the possibility of neonatal transmission via breast milk. Affected women may opt to defer vaccination or temporarily stop breast-feeding due to the lack of data on reproductive or lactational toxicity, even though COVID-19 vaccination is not contraindicated in pregnant or breastfeeding women, or for women planning pregnancy.^10, 11^ Given the enduring susceptibility of pregnant and peripartum women to COVID-19 complications, we find these trends worrying as they compound vaccine hesitancy among persons of reproductive age.^12^ Although attempts have been made to correct this misinformation by mainstream news outlets and the scientific community,^2, 13^ anti-vaccination pundits continue to propagate vaccine scepticism and dampen confidence in vaccine safety, even among female healthcare professionals.^14^ To begin to address these issues, we investigated post-vaccination mRNA persistence and anti-syncytin-1 antibodies in plasma and breast milk in female at-risk front-line workers and vaccine recipients.

## Methods

### Participants

Female front-line workers were approached at the National University Hospital, Singapore, to participate in this institutional review board-approved study (DSRB2012/00917) between February and April 2021. Volunteers were eligible if they were scheduled to receive BNT162B2 and consented to collection of blood and breast milk (where applicable) before and after the first dose. Data is presented according to STROBE guidelines.^15^

### Samples

Plasma was collected at Day 0 (pre-vaccination), 1-4 days and 4-7 weeks, while breast milk was collected within 24 hours and daily for the first week, following the first dose. Blood and expressed breast milk were centrifuged at 1500G and 3000G respectively. Plasma, aqueous and lipid factions of breast milk were flash frozen before total RNA extraction with Zymo Research Quick DNA/RNA viral kit (Zymo Research, Irvine, CA). Briefly, samples were treated with DNA/RNA shield, Proteinase K, and viral DNA/RNA buffer to bind DNA and total RNA to the Zymo-Spin™□ IIC-XLR Column. DNA was removed with DNase 1, RNA eluted in RNase-free Water, checked for purity and quantified spectrophotometrically.

### One-step real time quantitative PCR for detection of vaccine mRNA

TaqMan Primers were designed using the published mRNA sequence of BNT162B2 that was made available to the public,^16^ and amplified an 87bp segment of the spike-encoding region to determine the presence of vaccine mRNA in plasma and breast milk (Table 1). Isolated RNA, primers and probe were added to THUNDERBIRD™ Probe One-step (Toyobo, Osaka) qRT-PCR master mix and the real time PCR protocol was run on QuantStudio™ 1 Real-Time PCR System (Applied Biosystems, Foster City, CA). Negative controls were run simultaneously using water and RNA isolated from pre-vaccination plasma. Positive controls were day 1 post-vaccination RNA samples.

**Table 1.**
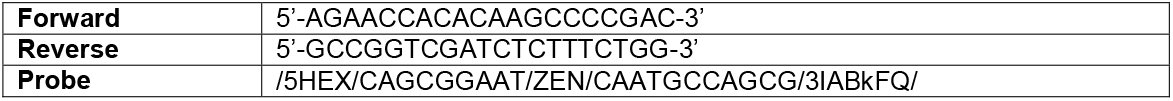
Primers used to amplify a region of the spike-encoding region of the BNT162B2 mRNA.

### Library preparation and whole transcriptome sequencing

First and second strand synthesis was performed with extracted plasma RNA as input (Ovation® RNA-seq system v2 kit, NuGEN Technologies Inc, Redwood City, CA). Illumina Nextera XT DNA Library Preparation kit was used for library production and barcoding. Paired end-sequencing of 300bp read length was performed on the iSeq100 sequencer (Illumina, San Diego, CA).

### Sequenced read analyses and assembly

Sequenced reads were aligned to human references using HISAT2 and filtered out. Remaining non-human reads were assembled using SPAdes. Comparative analyses with published Pfizer sequences and MT380725 were performed with MUMMER. Multiple genome alignment and visualization were done using the MUSCLE and R package ggmsa (https://github.com/YuLab-SMU/ggmsa). ^17-20^

### Anti-syncytin-1 antibody semi-quantitative ELISA

96-well Maxisorp plates (Thermo Fisher Scientific, Massachusetts) were coated with 100ng of human syncytin-1 recombinant protein (MyBioSource, San Diego) overnight (4°C), then washed and blocked with 6% bovine serum albumin (BSA; Sigma, Missouri) in 0.05% Tween:1X PBS (PBST) at 37°C. Pre-vaccination plasma served as the negative control, giving an optical density reading at an absorbance wavelength of 492nm (OD_492_) of ∼0.1.

Positive controls were produced by coating wells with 100ng of human syncytin-1 recombinant protein and incubating with rabbit anti-human syncytin-1 antibody (MyBioSource) at 1:250 dilution, selected after optimization as it gave a consistent OD_492_ of ∼0.9. This readout was based on standard ELISAs, as there is no data on clinically-significant thresholds of anti-syncytin-1 antibodies.

Samples and controls were diluted 1:50 with dilution buffer (2% BSA in PBST), incubated in prepared plates at room temperature, then incubated with horseradish peroxidase-conjugated goat anti-rabbit secondary antibody (for positive controls, at 1:1000 dilution; Thermo Fisher Scientific, Massachusetts), or goat anti-rhesus secondary antibody (for all samples, at 1:4000 dilutions; Southern Biotech, Alabama), washing between steps. Plates were loaded with o-phenylenediamine dihydrochloride substrate (Sigma) and colorimetric reactions arrested at 3 minutes with 3M HCl. OD_492_ was analysed in the Sunrise microplate reader (Tecan, Mannedorf, Switzerland). Background absorbance was eliminated with a blank control of dilution buffer.

### SARS-COV-2 Neutralising Antibody assay

Neutralizing antibodies (NAb) against SARS-CoV-2 were detected using the SARS-CoV-2 Surrogate Virus Neutralization Test Kit (GenScript, NJ). Samples and controls were pre-incubated with HRP-Receptor Binding Domain (RBD) to allow the binding of circulating NAb to HRP-RBD, and the mixture added to the capture plate (pre-coated with human angiotensin converting enzyme-2 protein). Circulating NAb HRP-RBD complexes in the supernatant, if any, were removed during washing while unbound HRP-RBD and HRP-RBD bound to non-neutralizing antibodies remained on the capture plate. Colorimetric reactions were elicited and stopped with the addition of TMB and Stop solutions respectively, and colour intensity read at absorbance wavelength of 450nm in the Sunrise microplate reader. Inhibition was calculated based on the OD_450_ absorbance (inversely proportionate to anti-SARS-CoV-2 NAb titers) and inhibition ≥30% was interpreted as a positive result according to manufacturer’s instructions.

### Post-hoc analysis

Results are expressed as mean±SD and compared by one-way ANOVA using Tukey correction for multiple comparisons.

## Results

### Participants

Fifteen female at-risk front-line workers who were eligible for the vaccine, including five breast-feeding mothers and two women inadvertently vaccinated in early pregnancy, were recruited for this institutional review board-approved study (DSRB2012/00917) conducted at the National University Hospital of Singapore. No subject was previously diagnosed with COVID-19. Mean age was 40.4±12.2 years. Participants were of Malay, Indian and Chinese ethnicities. All women received two doses of BNT162B2 according to the prescribed schedule, except the two pregnant subjects who each had a single dose before pregnancy confirmation, and subsequently did not receive the second dose (Table 2).

**Table 2.** Participant characteristics.

|  | **No. (%)** |
| --- | --- |
| **Study participants** |  |
| Approached to participate in study | 18 |
| Consented, completed study | 15 (83.3) |
| Consented, did not complete study (vaccination deferred) | 1 (5.6) |
| Declined participation | 2 (11.1) |
| **Participant features** | 15 (100.0) |
| Age, mean (SD), y | 40.4 (12.2) |
| Parous | 14 (93.3) |
| Pregnant at vaccination | 2 (13.3) |
| Received 2 doses | 13 (86.7) |
| Received 1 dose | 2 (13.3) |
| Malay ethnicity | 4 (26.7) |
| Indian ethnicity | 4 (26.7) |
| Chinese ethnicity | 7 (46.6) |
| **Samples collected** |  |
| Day 0 plasma | 13/15 (86.7) |
| Day 1-4 plasma | 13/15 (86.7) |
| Week 4-5 plasma | 8/15 (53.3) |
| Week 6-7 plasma | 6/15 (40.0) |
| Day 0-7 breast milk | 5/5 (100.0) |
| **Incomplete samples** |  |
| Unable to return for blood collection (late time-point) | 1 (6.7) |
| Insufficient plasma for analysis (early time-point) | 1 (6.7) |

### Vaccine mRNA was amplified from post-vaccination plasma and perfectly aligned with the published sequence

Plasma BNT162B2 mRNA was detected within 4 days of vaccination (n=13), including in all breastfeeding women (Ct<30, all samples, Figure 1A,B), but no amplification was observed in either aqueous or lipid breast-milk factions (days 0-7, n=5, Figure 1B). Early plasma samples were not obtained from the two participants with undiagnosed pregnancies at the time of vaccination as they had not yet been recruited (subjects 101, 102); subsequently these two participants did not amplify plasma BNT162B2 mRNA at week 4, the only single-dose samples at this time point as the other recipients had received their second dose by week 3.

**Figure 1.**
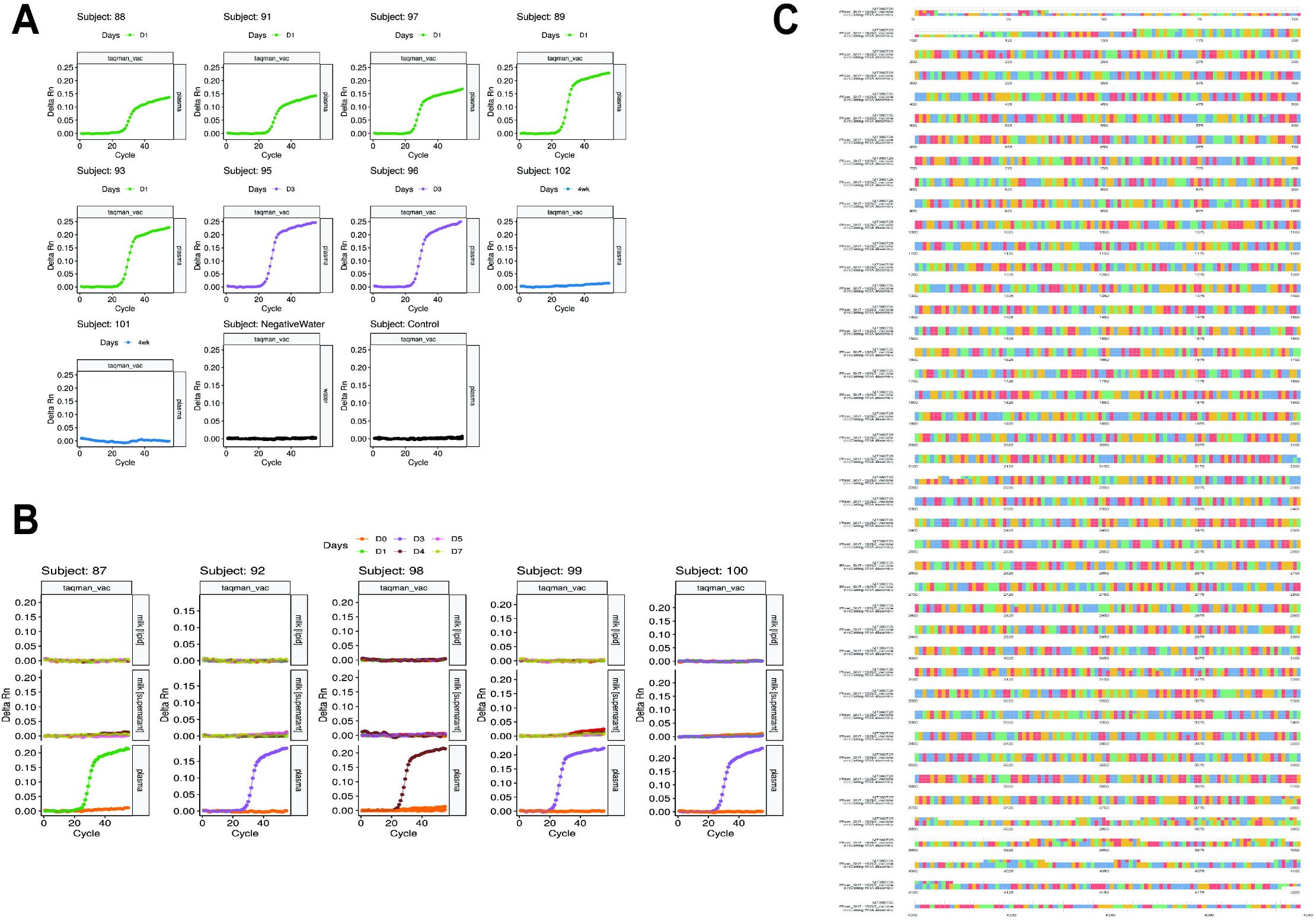
Detection of BNT162B2 mRNA and Multiple sequence alignment of plasma RNA vaccine assembly, Pfizer sequence and MT380725 spike protein sequence. (A) Amplification at Ct<30 was observed in all plasma samples between 1 and 4 days post-vaccination, but not in 4-week plasma collected from subjects 101 and 102 (undiagnosed pregnancies at the time of the first dose). Negative control (pre-vaccination plasma) and water blanks did not amplify, demonstrating the specificity of the TaqMan primers/probe. (B) Five paired breast milk and plasma samples were analysed for BNT162B2 mRNA, which amplified (Ct<30) in day 1-4 plasma but not in day 0-7 breast milk. Negative control (pre-vaccination plasma) and water blanks did not amplify, demonstrating the specificity of the TaqMan primers/probe. (C) An 87bp region of the spike protein-encoding region of the BNT162B2 mRNA was amplified from RNA extracted from plasma. Whole transcriptome sequencing was performed on extracted RNA. A draft scaffold of 4196 bases with an average coverage of 162X, of comparable size to the vaccine RNA, was obtained only from non-human sequenced reads of post-vaccination samples. No scaffolds >2kb were obtained from pre-vaccination plasma. We observed perfect alignment from bases 120 to 4077 when we compared our assembled sequence to the public Pfizer sequence, and a high degree of agreement at the nucleotide level (other than at the start and end of the sequences) when multiple sequence alignment was performed with GenBank sequence MT380725 of the SARS-CoV-2 spike protein.

High-quality non-human sequenced reads from whole transcriptome sequencing of plasma- and breast milk-extracted circulating RNA were assembled to obtain draft genomic scaffolds of 4196bp (average coverage of 162X), comparable in size to BNT162B2 mRNA, and we observed perfect alignment from 120-4077bp against the published sequence. No scaffolds greater than 2kb were obtained for pre-vaccination plasma. Multiple sequence alignment performed with GenBank sequence MT380725 (SARS-CoV-2 spike protein) demonstrated agreement at the nucleotide level (Figure 1C).

### Anti-syncytin antibodies were not detected in post-vaccination plasma

To determine if recipients mounted the desired immune response to BNT162B2, we first analysed SARS-CoV-2 neutralising antibodies (NAb), finding negative responses at day 0-4 (post-vaccination inhibition at 14.7±4.7%), and strongly positive responses 4-7 weeks post-vaccination (inhibition at 98.7±1.2%, p<0.001, Figure 2A), including in the pregnant women (inhibition >89.0% following a single dose). At the same time-points, anti-syncytin-1 binding activities were far below the positive control and were interpreted as negative (Figure 2B).

**Figure 2.**
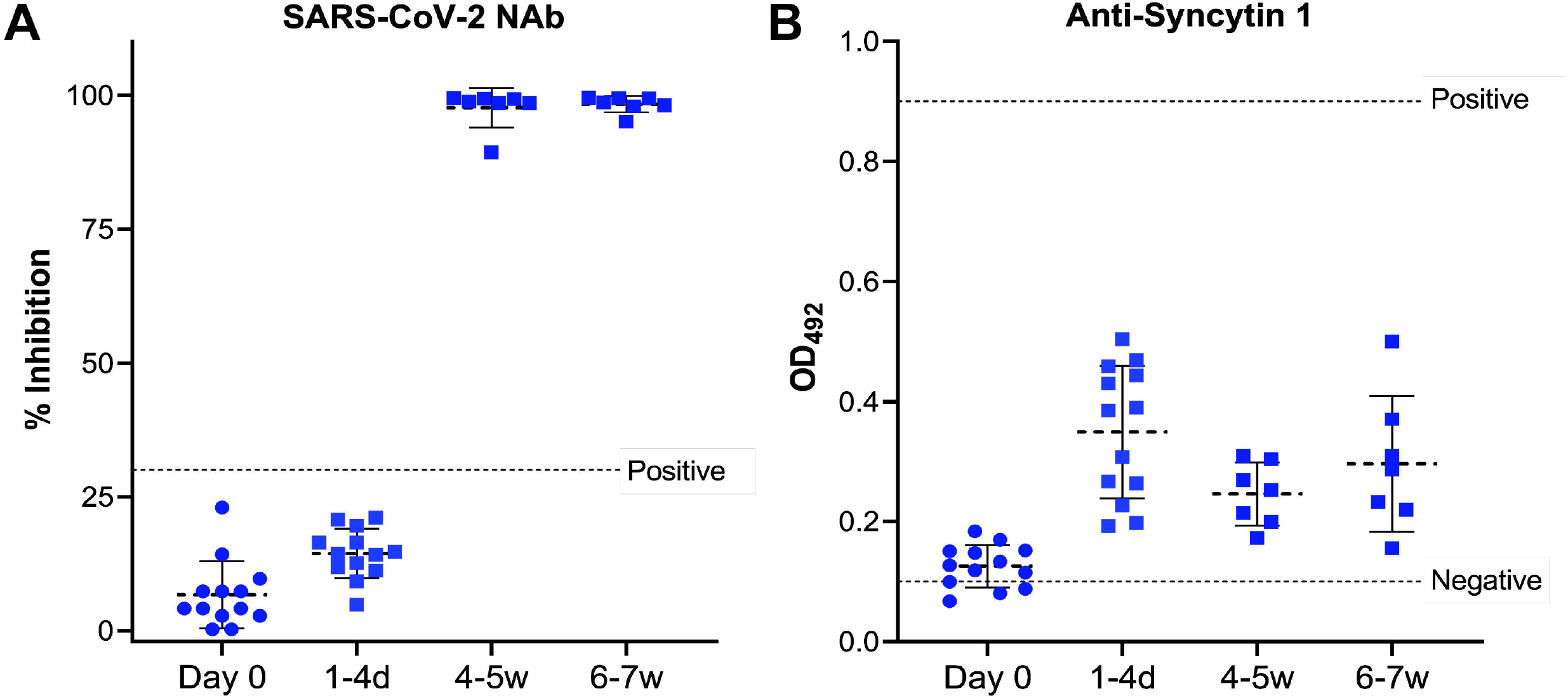
Antibodies to SARS-CoV-2 and syncytin-1. A commercial neutralising assay was used to detect SARS-CoV-2 neutralising antibodies, and an ELISA was designed in-house to determine the presence of anti-syncytin-1 antibodies. (A) All subjects were negative for SARS-CoV-2 neutralising antibodies on days 0-4 and strongly positive by at least week 4, and none showed co-existing binding antibodies to human syncytin-1 antigen (B). Dashed line represents mean, error bars represent SD; d-days, w-weeks.

## Discussion

Our study shows that BNT162B2-vaccinated women did not transmit vaccine mRNA to breast milk, and supports the continuation of breastfeeding which may transfer protective antibodies.^21^ These data are limited by the lack of surveillance beyond one week post-vaccination. Vaccinated woman should be informed about the current absence of breastfeeding safety data.^10^ Our small study is also the first to investigate anti-syncytin antibodies following BNT162B2 vaccination. Our finding that BNT162B2-vaccinated women did not produce a significant antibody response to syncytin-1, despite demonstrating strong neutralising activity against SARS-CoV-2, is important because it suggests that cross-reactivity to syncytin proteins on the developing trophoblast is unlikely. This is in line with computerised basic local alignment search tool (BLAST) comparisons of published nucleotide and protein sequences which have shown very limited amino acid sequence similarity – only two 2-amino acid stretches – between syncytin-1 and the S2 domain of the SARS-CoV-2 spike protein; at least 80 continuous amino acid stretches comprising >35% sequence homology is required for cross-reactivity between proteins to occur.^4^ We did not however, examine duration of mRNA persistence or the clinical significance of anti-syncytin-1 antibody levels. As such, we encourage a restrained interpretation of our findings, as post-authorisation surveillance data from the US Vaccine Adverse Event Reporting System (VAERS) highlight spontaneous miscarriage as the most common obstetric outcome after COVID-19 mRNA vaccination.^22^ Longitudinal surveillance of larger numbers of reproductive-age women vaccinated pre-conception and in early pregnancy should assist in parsing the role of anti-syncytin antibodies in fertility and pregnancy.

## Data Availability

Data will be made available by the authors upon request

## Acknowledgements

This study was funded by National University of Singapore IRB Appointment Research Fund to PAT and National Medical Research Council, Singapore COVID-19 Gap Funding (COVID19FR3-0090) to WK,YS, SH.

## References

1. S. Stecklow and A. MacAskill. SPECIAL REPORT The ex-Pfizer scientist who became an anti-vax hero. In Book SPECIAL REPORT The ex-Pfizer scientist who became an anti-vax hero, Editor (ed)^(eds). Reuters: City, 2021.

2. V. Male. Are COVID-19 vaccines safe in pregnancy? Nat Rev Immunol 2021; 21: 200–201. DOI 10.1038/s41577-021-00525-y.

3. L. Hamel, A. Kirzinger, L. Lopes, A. Kearney, G. Sparks and M. Brodie. KFF COVID-19 Vaccine Monitor: January 2021. Vaccine Hesitancy. In Book KFF COVID-19 Vaccine Monitor: January 2021. Vaccine Hesitancy., Editor (ed)^(eds). KFF: City, 2021.

4. B. Bjerregaard, J. G. Lemmen, M. R. Petersen, E. Østrup, L. H. Iversen, K. Almstrup, L. I. Larsson and S. Ziebe. Syncytin-1 and its receptor is present in human gametes. Journal of Assisted Reproduction and Genetics 2014; 31: 533–539. DOI 10.1007/s10815-014-0224-1.

5. R. M. Roberts, T. Ezashi, L. C. Schulz, J. Sugimoto, D. J. Schust, T. Khan and J. Zhou. Syncytins expressed in human placental trophoblast. Placenta 2021. DOI 10.1016/j.placenta.2021.01.006.

6. B. Soygur and H. Moore. Expression of Syncytin 1 (HERV-W), in the preimplantation human blastocyst, embryonic stem cells and trophoblast cells derived in vitro. Hum Reprod 2016; 31: 1455–1461. DOI 10.1093/humrep/dew097.

7. B. S. Holder, C. L. Tower, V. M. Abrahams and J. D. Aplin. Syncytin 1 in the human placenta. Placenta 2012; 33: 460–466. DOI 10.1016/j.placenta.2012.02.012.

8. B. Soygur, L. Sati and R. Demir. Altered expression of human endogenous retroviruses syncytin-1, syncytin-2 and their receptors in human normal and gestational diabetic placenta. Histology and histopathology 2016; 31: 1037–1047. DOI 10.14670/hh-11-735.

9. M. Kloc, A. Uosef, J. Z. Kubiak and R. M. Ghobrial. Exaptation of Retroviral Syncytin for Development of Syncytialized Placenta, Its Limited Homology to the SARS-CoV-2 Spike Protein and Arguments against Disturbing Narrative in the Context of COVID-19 Vaccination. Biology (Basel) 2021; 10. DOI 10.3390/biology10030238.

10. Centers_for_Disease_Control_and_Prevention. Information about COVID-19 Vaccines for People who Are Pregnant or Breastfeeding. In Book Information about COVID-19 Vaccines for People who Are Pregnant or Breastfeeding, Editor (ed)^(eds). CDC: City, 2021.

11. Ministry_of_Health_Singapore. Vaccination Information Sheet (for Vaccination Recipients). In Book Vaccination Information Sheet (for Vaccination Recipients), Editor (ed)^(eds). Ministry of Health, Singapore: City, 2020.

12. S. Kadiwar, J. J. Smith, S. Ledot, M. Johnson, P. Bianchi, N. Singh, C. Montanaro, M. Gatzoulis, N. Shah and E.-F. Ukor. Were pregnant women more affected by COVID-19 in the second wave of the pandemic? The Lancet 2021. DOI 10.1016/s0140-6736(21)00716-9.

13. A. Rodriguez. No, the COVID-19 vaccine doesn’t cause infertility in women. In Book No, the COVID-19 vaccine doesn’t cause infertility in women, Editor (ed)^(eds). USA TODAY: City, 2020.

14. Pulse on the Nation’s Nurses COVID-19 Survey Series: COVID-19 Vaccine. https://www.nursingworld.org/practice-policy/work-environment/health-safety/disaster-preparedness/coronavirus/what-you-need-to-know/covid-19-vaccine-survey/.

15. E. von Elm, D. G. Altman, M. Egger, S. J. Pocock, P.C. Gøtzsche and J. P. Vandenbroucke. The Strengthening the Reporting of Observational Studies in Epidemiology (STROBE) Statement: guidelines for reporting observational studies. Int J Surg 2014; 12: 1495–1499. DOI 10.1016/j.ijsu.2014.07.013.

16. F. P. Polack, S. J. Thomas, N. Kitchin, J. Absalon, A. Gurtman, S. Lockhart, J. L. Perez, G. Pérez Marc, E. D. Moreira, C. Zerbini, R. Bailey, K. A. Swanson, S. Roychoudhury, K. Koury, P. Li, W. V. Kalina, D. Cooper, R. W. Frenck Jr.,, L. L. Hammitt, Ö. Türeci, H. Nell, A. Schaefer, S. Ünal, D. B. Tresnan, S. Mather, P. R. Dormitzer, U. Şahin, K. U. Jansen and W. C. Gruber. Safety and Efficacy of the BNT162b2 mRNA Covid-19 Vaccine. The New England journal of medicine 2020; 383: 2603–2615. DOI 10.1056/NEJMoa2034577.

17. D. Kim, J. M. Paggi, C. Park, C. Bennett and S. L. Salzberg. Graph-based genome alignment and genotyping with HISAT2 and HISAT-genotype. Nat Biotechnol 2019; 37: 907–915. DOI 10.1038/s41587-019-0201-4.

18. R. C. Edgar. MUSCLE: a multiple sequence alignment method with reduced time and space complexity. BMC Bioinformatics 2004; 5: 113. DOI 10.1186/1471-2105-5-113.

19. A. Bankevich, S. Nurk, D. Antipov, A. A. Gurevich, M. Dvorkin, A. S. Kulikov, V. M. Lesin, S. I. Nikolenko, S. Pham, A. D. Prjibelski, A. V. Pyshkin, A. V. Sirotkin, N. Vyahhi, G. Tesler, M. A. Alekseyev and P. A. Pevzner. SPAdes: a new genome assembly algorithm and its applications to single-cell sequencing. J Comput Biol 2012; 19: 455–477. DOI 10.1089/cmb.2012.0021.

20. A. L. Delcher, A. Phillippy, J. Carlton and S. L. Salzberg. Fast algorithms for large-scale genome alignment and comparison. Nucleic Acids Res 2002; 30: 2478–2483. DOI 10.1093/nar/30.11.2478.

21. S. H. Perl, A. Uzan-Yulzari, H. Klainer, L. Asiskovich, M. Youngster, E. Rinott and I. Youngster. SARS-CoV-2–Specific Antibodies in Breast Milk After COVID-19 Vaccination of Breastfeeding Women. JAMA 2021. DOI 10.1001/jama.2021.5782.

22. T. T. Shimabukuro, S. Y. Kim, T. R. Myers, P. L. Moro, T. Oduyebo, L. Panagiotakopoulos, P. L. Marquez, C. K. Olson, R. Liu, K. T. Chang, S. R. Ellington, V. K. Burkel, A. N. Smoots, C. J. Green, C. Licata, B. C. Zhang, M. Alimchandani, A. Mba-Jonas, S. W. Martin, J. M. Gee, D. M. Meaney-Delman and C. D. C. v.-s. C.-P. R. Team. Preliminary Findings of mRNA Covid-19 Vaccine Safety in Pregnant Persons. The New England journal of medicine 2021. DOI 10.1056/NEJMoa2104983.

